# A Behavioral Multispread Epidemic Model

**DOI:** 10.1101/2020.08.24.20181107

**Authors:** Carlo Graziani

## Abstract

We introduce a class of epidemic models that represent multiple spread rates in terms of discrete behavior classes, rather than in terms of discrete compartments comprising individuals. The model is framed in terms of *D* behavior classes, each with its own spread rate. The population is represented as a density on the *D*-simplex, where each point is a *D*-vector ***f*** whose components sum to 1. Each component of ***f*** represents the fraction of time in which an individual spends engaging in the corresponding behavior. The evolution equation is an integro-differential equation on the *D*-simplex. The model is capable of describing the “superspreader” phenomenon in terms of behavior spread rates, as opposed to terms of individual infectivity. We show the existence of SIR-like separable solutions and discuss their stability. We explore the numeric properties of the model using a *D* = 3 case featuring a “safe” behavior, a moderate-spread behavior, and a superspread behavior.

## 1. Introduction

The COVID-19 global epidemic has brought renewed focus on heterogeneous spread mechanisms and specifically on the “superspreader” phenomenon [see, e.g., 1, 2, 3]. In qualitative epidemiological discussions, the term “superspreader” is used in several distinct senses, which can often be found commingled in discussions of the phenomenon. In one sense, a “superspreader” is an infected individual whose physiology results in higher infectivity than normal. Another sense of the term refers to events, rather than to individuals. In the context of the COVID-19 epidemic, superspreader events such as weddings, funerals, or political rallies have been frequently cited as likely accelerators of the epidemic. “Superspreader” may also refer to residential or geographic circumstances such as dense urban areas or assisted-care facilities, where spread is observed to be more rapid than elsewhere. Moreover, the term may refer to behaviors such as frequent attendance at bars, restaurants, and religious services. Obviously, some overlap between these categories exists and explains their frequent conflation in discussions of the phenomenon.

In technical/mathematical epidemiology the first sense of the term appears to dominate modeling. Efforts to model multiple spread rates through systems of ordinary differential equations (ODEs) [4], partial differential equations [5, 6, Chap. 12], delay-differential equations [7], stochastic differential equations [8], or integro-differential equations [9] rely on the first, individual-physiological notion of infectivity, because such models borrow the compartmental structure of classic epidemiological models such as the SIR model of Kermack and McKendrick [10], with discrete compartments representing groupings of susceptible, exposed, infected, and recovered/removed individuals. Dynamically speaking, in these models the compartments are fields over time, or over space-time. Such models differ in compartment selection and in dynamical rules for transferring individuals between compartments, but the role and interpretation of the compartments themselves are invariant.

Efforts have been made to model superspreader *events* (as opposed to individuals) that do not change the dynamical stage of compartment fields over time and possibly space and that rely on stochastic source terms to model high-spread events [11,12]. While these efforts model spread variability by using stochastic processes, they do not appear to explore behavior explicitly. Herein is a further subtle distinction in the use of “superspreader”: we think of the term differently when we refer to “superspreader events” and“superespreader behavior.” To speak of a set of “events” evokes, in the mind of a modeler, a stochastic process, which is readily superposed on the existing dynamical infrastructure. A set of “behaviors,” on the other hand, calls to mind stable, deterministic relations and leads more naturally to a replacement of the dynamical stage of time, or space-time, with a new stage that we refer to below as “behavior space.”

We are interested in exploring the possible structure of a deterministic epidemic model embodying the second meaning of the term “superspreader”: a model in which behaviors—rather than specific categories of individuals—are assigned different infectivities, and individuals become infected to the degree that they participate in such activities. We present one such model here: the *behavioral multispread* (BMS) model.

The plan of this article is as follows. In Section §2 we introduce the BMS model equations and discuss their interpretation. In §3 we derive separable solutions of the BMS equations. In §4 we discuss the stability of the infection-free solutions and of the separable solutions, discuss the nature of herd immunity in the BMS model, and consider late-time asymptotic behavior. In §5 we illustrate the nature of the BMS model in the context of a 3-behavior model featuring a very conservative (i.e., nonpropagating) behavior, a marginally propagating behavior, and a superspread behavior. A discussion and summary follows in §6. Some technical theorems concerning local and global stability of the BMS equations are proven in Appendix A and Appendix B.

## 2. The Model

### 2.1. Framework

The BMS model assumes a population characterized by the degree of participation in each of D behaviors or activities. An individual is characterized by a vector ***f*** on the D-simplex, that is, satisfying

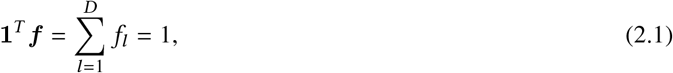

and *f_l_* ≥ 0, *l =* 1,…, D. Each component *f_l_* is interpreted as the fraction of time spent by the individual at the *l*th behavior. The simplex condition enforces the condition that all of the individual’s time is accounted for. In Equation (2.1) we have introduced the “one” vector **1** whose components are all equal to 1.

Each behavior is characterized by its own infectivity, which we parametrize by a parameter *a_l_*, *l* = 1,…, *D*. The meaning of these parameters and their relation to epidemiological reproduction numbers will be made clear below. It is convenient to define the diagonal matrix ***A*** ≡ Diag(*a*_1_,…, *a_D_*).

We require a normalized population density on the *D*-simplex. For this purpose we introduce a (*D* − 1)-dimensional measure *dη*(***f***), which we may conveniently define by 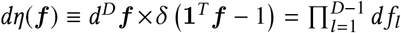, where *δ*(*x*) is the Dirac point-mass distribution “density.” We define a population density *ρ*(*f*) and measure *dμ*(***f***) ≡ *ρ*(***f***)*dη*(***f***), satisfying 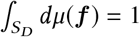, where *S_D_* is the *D*-simplex. With these definitions, we may easily choose a standard form for *ρ*(***f***) such as a member of the family of Dirichlet distributions, or a mixture thereof. Alternative and more general distributions on the simplex with well-understood properties are known from the theory of statistical analysis of compositional data [13]. In the BMS model the measure *dμ*(***f***) is not dynamical but, rather, is regarded as part of the fixed structure of the model. The kinematic structure of the BMS model is establishedd by fixing the triplet (*S_D_*, ***A***, *dμ*(***f***)).

The dynamical structure of the BMS model is defined in terms of the local susceptible, infected, and removed fractions *s*(*t*, ***f***), *i*(*t*, ***f***), and *r*(*t*, ***f***), respectively, satisfying *s* + *i* + *r* = 1. These functions are defined in a manner similar to the corresponding fractions from the well-known SIR compartmental model [10,6]. Their interpretation is that *s*(*t*, ***f***) is the fraction of the population characterized by the simplex point ***f*** that is susceptible to infection, and similarly for *i*(*t*, ***f***) and *r*(*t*, ***f***). The total susceptible and infected compartment fractions in the population are thus *S*(*t*) = ∫ *dμ*(***f***) *s*(*t*, *f*) and *I*(*t*) = ∫ *dμ*(***f***) *i*(*t*, *f*), with the removed fraction being simply R = 1 − *S* − *I* as usual.

We assume that individuals are removed at a uniform rate *ν* from the infected state at each ***f***. The parameter *ν* furnishes a natural time scale, so we will use that scale as a unit of time, defining the dimensionless time variable *τ* ≡ *νt*

### 2.2. Dynamical System

The model dynamics for the fractions i and s are given by the following equations

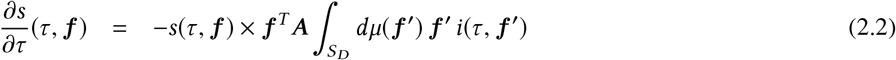

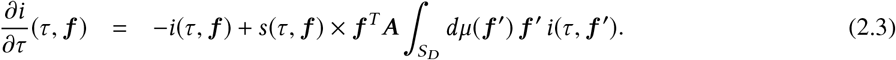

These equations are inspired by the SIR model with demography neglected, so that natural births and deaths not considered. Equations (2.2-2.3) are of easy interpretation. Equation (2.2) states that susceptibles at the point ***f*** ∊ *S_D_* are infected by contact with infected individuals at other points ***f′*** ∊ *S_D_* at a rate equal to 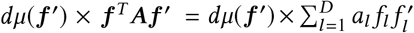, which is to say at a sum of rates, the *l*th of which is in proportion to the respective fractions spent in behavior *l* by the susceptible individuals indexed by ***f*** and by the infected individuals indexed by ***f′***. Equation (2.3) states that all infections of susceptibles at ***f*** result in the creation of infected individuals at ***f*** and that the fraction of such infected individuals decays at the removal rate, which is 1 with respect to the dimensionless time *τ*.

The parameter *a_l_* plays a role analogous to a reproduction number for behavior *l*. It will be larger for riskier behaviors than for more conservative behaviors. By integrating Equations (2.2-2.3) with respect to *dμ*(***f***) we may define a time-dependent “effective reproduction number” *R_E_* (*τ*) for the system:

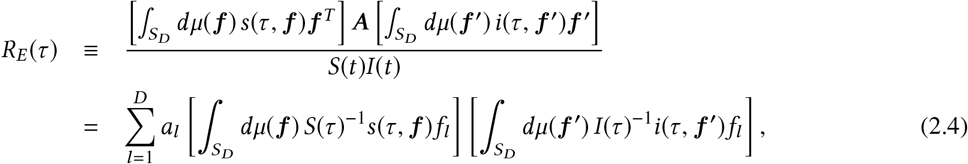

in terms of which we have the SIR-like effective equations

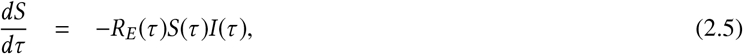

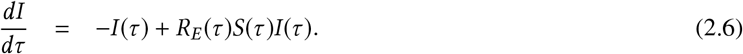

These equations require the solution of the full system of Equations (2.2-2.3) to obtain *R_E_* (*τ*) and hence are not practical tools for obtaining such solutions. They are nonetheless helpful, illustrating the relationship of the BMS model to the SIR model. To gain intuition concerning the relation of the parameters *a_l_*; and *R_E_*(*τ*), suppose that both the weighted averages

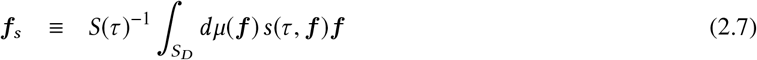

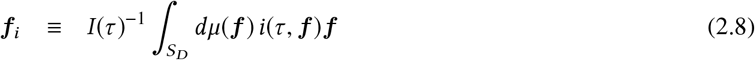

are located near the simplex uniform mean point, ***f****_s_* ≈ ***f****_i_* ≈ *D*^−1^**1**. In this case we would obtain *R_E_*(*τ*) ≈ *D*^−2^Tr***A*** = *D*^−1^*ā*, where *ā* is the unweighted average of the *a_l_*.

In §3 we show the existence of true SIR-type separable solutions of the BMS equations, and we establish the relationship between the parameters al and the basic reproduction number of those solutions. This will be convenient since the resulting relation will not depend on specific forms of the functions *i*(·), *f*(·).

Equations (2.2–2.3) may be solved numerically by discretizing the simplex *S_D_* and writing the resulting system as a (high-dimensional) ODE. In §5 we demonstrate this method of solution in a *D* = 3 system using a triangularizing discretization of the simplex.

## 3. Separable Solutions

We assume the existence of separable solutions of the form

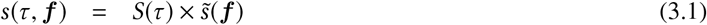

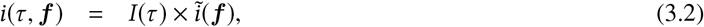

where the functions 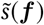 and *ĩ*(***f***) are normalized:

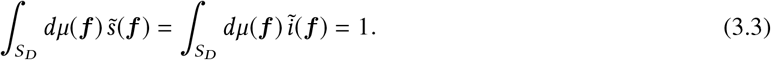

Substituting these into Equation (2.2), we obtain

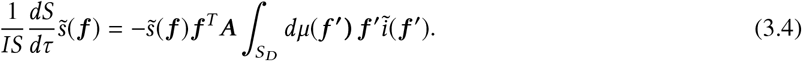

Since the right-hand side of Equation (3.4) is independent of *τ*, we must have

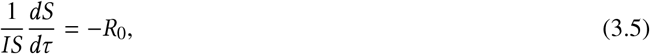

where *R*_0_ is a constant independent of *τ*. We may use this to rewrite Equation (2.3) as follows:

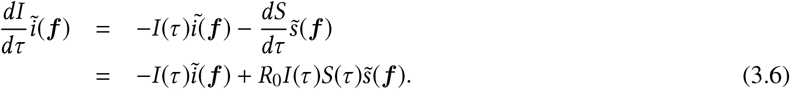

This establishes that the functions 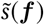 and *ĩ*(***f***) are proportional to each other: 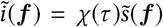. However, by the normalization condition of Equation (3.3) it follows that *χ*(*τ*) = 1, and hence

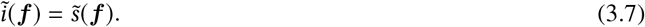

Equations (3.5) and (3.6) now reduce to

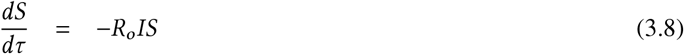

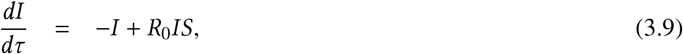

which is just the SIR system of equations, with *R*_0_ recognizable as the basic reproduction number.

We now combine Equations (3.4), (3.5), and (3.7) to obtain

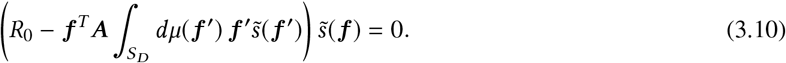

Since this equation holds for all ***f*** ∊ *S_D_*, it follows that 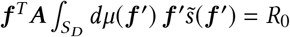 for all ***f*** *∊ S_D_*, which is possible only if

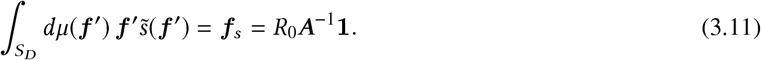

Since 1*^T^* ***f*** = 1, we obtain the relation

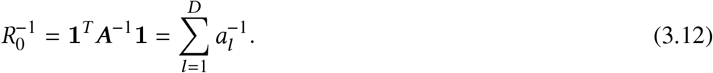

Using Equations (2.4), (3.1), (3.2), (3.11), and (3.12), one can easiliy demonstrate that for a separable solution *R_E_* (*τ*) = *R*_0_.

Thus, 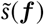 may not be chosen freely but must satisfy an integral constraint on the value of ***f****_s_* given by Equations (3.11) and (3.12) in terms of the parameters *a_l_*. The nature of the constraint is that ***f****_s_* —the centroid on *S_D_* of the measure 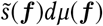–has components *f_sl_* that are inversely proportional to the *l*-th infectivity parameter *a_l_*. Thus the more infective the *l*th behavior, the smaller the concentration of 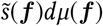 and of *ĩ*(***f***)*dμ*(***f***) in regions of *S_D_* where ***f*** ≈ 1, and hence the greater the concentration of removed individuals 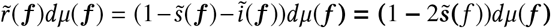 in such regions. We may interpret this observation if we may regard a separable solution as an asymptotic limit of a solution started at an arbitrary state—say, almost 100% of the population susceptible except for a small infected population at ***f***_0_. Then we may say that the most infective regions oUsing Equations (2.4), (3.1), (3.2), (3.11), and (3.12), one can easiliy demonstrate that for a separable solution *R_E_*(*τ*) = *R*_0_. f *S_D_* are the ones most thoroughly emptied of susceptibles, who are replaced by removed/recovered individuals.

In order for such an interpretation to be tenable, the stability of asymptotic solutions of the BMS equations must be studied. The required analysis follows in §4. It will show that general solutions of the BMS equations in fact are initially attracted towards separable solutions. However separable solutions are not globally stable, whereas infection-free solutions are globally stable. As a consequence, initial convergence towards a separable solution is cut off by the advent of convergence towards an infection-free solution, a phenomenon we will refer to as *freeze-out* below.

## 4. Stability of Solutions

### 4.1. Stability of the Infection-Free Solution

Recall that the SIR equations (3.8) and (3.9) have an infection-free solution *S* = *S*_0_, *I* = 0, which is globally stable if *S*_0_*R*_0_ ≤ 1 (herd immunity) and unstable otherwise [6, Chap. 2].

The BMS equations have an analogous special solution in which 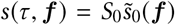, *i*(*τ*, ***f***) = 0, where the function 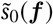 is normalized as in Equation (3.3). This solution is stable only under certain conditions that we now derive. Linearizing by

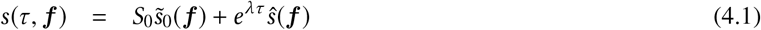

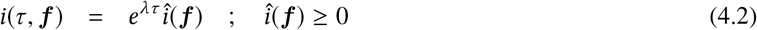

and substituting in Equations (2.2) and (2.3), we find to linear order

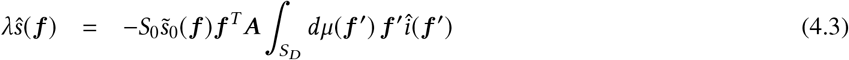

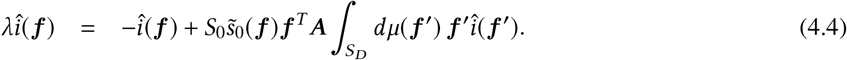

Adding these equations, we immediately obtain 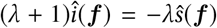. The condition *î*(***f***) > 0 rules out nontrivial solutions for both *λ* = 0 and *λ* + 1 = 0. We may write

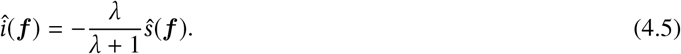

Combining this with Equation (4.3), we find

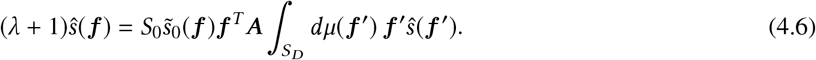

It is apparent from Equation (4.6) that the dependence on f of *ŝ*(***f***) requires that there exist an ***f*** -independent *D*-vector *q_λ_* such that

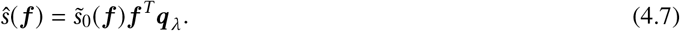

Inserting this back in Equation (4.6) results in

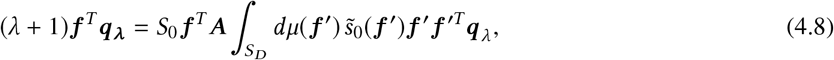

which can hold for all f only if *q_λ_* is an eigenvector of the operator on the RHS, that is, if

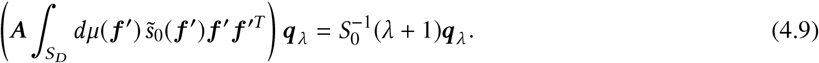

Since 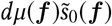 is a probability measure, we may appeal to Theorem 2 in Appendix Appendix A. Defining the mean vector 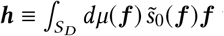 with components *h_l_*, and noting that in general ***h*** ≠ *R*_0_***A***^−1^**1** (that is, Equation 3.11 is not required to hold), by Theorem 2 we have that *λ* is real and that

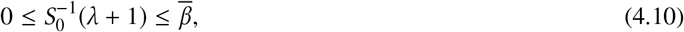

where we have defined the bound 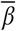 as

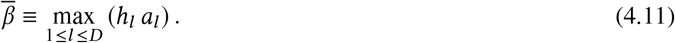

From Equation (4.10) we see that

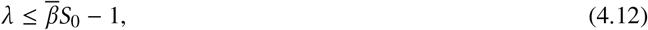

and that the herd immunity condition *λ* < 0 can always be achieved by making *S*_0_ sufficiently small that

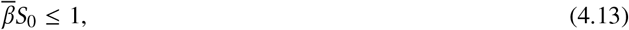

similarly to the herd immunity condition of the SIR model, but with 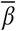 in the role of *R*_0_. This is now a statement not only about the value of *S*_0_ but also (through 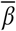) about the distribution density 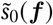 of susceptibles on *S_D_*. If there is no epidemic and if susceptibles are arranged so that their mean ***h*** has components such that 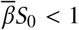, a new outbreak will not cause a propagating epidemic.

In the context of an ongoing epidemic, we may view 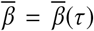 as a function of time, constructed from the current normalized state 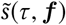, that is,

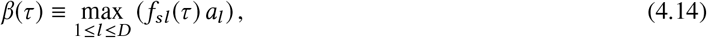

where *f_sl_* (*τ*) is the *I*-th component of the moment ***f****_s_* (*τ*) defined in Equation (2.7). By Theorem 3 of Appendix Appendix B we have the herd immunity condition for an in-progress epidemic:

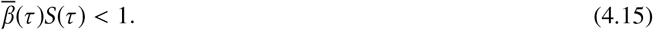

If Equation (4.15) is satisfied, then by Theorem 3 the solution is guaranteed to tend to an infection-free solution *i* = 0 as *τ* → ∞.

Thus the BMS herd immunity condition has meaning both as a local and a global stability condition, analogously to the herd immunity condition of the SIR model [6].

### 4.2. Stability of Separable Solutions

The local stability analysis of the separable solutions derived in §3 is similar to that of the infection-free solution. We start with the linearization

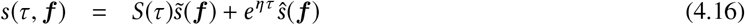

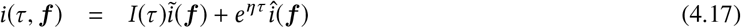

which perturbs the solutions described by Equations (3.1-3.2) by 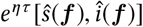.

Since we are perturbing about a time-dependent solution, the linearized perturbation equations contain time-dependent parameters, and we anticipate obtaining eigenvalues *η*(*t*) that are time-dependent. It is therefore necessary to impose a scale-separation condition on the perturbations, to ensure that the results make sense. In particular, the time-dependence of the perturbations is *e^η^*^(^*^τ^*^)^*^τ^*, which can only be interpreted as an exponential decay or growth if *dη*/*dτ* is in some sense small.” To make this precise, we Taylor-expand the exponential at time *τ*_0_. At time *τ* = *τ*_0_ + Δ*τ*, the perturbation time-dependence is

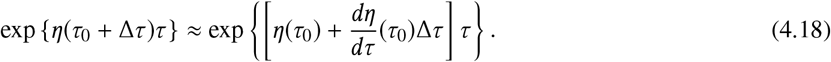

In order for the exponential decay (or growth) not to be spoiled, the second term in the exponential of Equation (4.18) must be small compared to the first over the course of an exponentiation time Δ*τ* = *η*(*τ*_0_)^−1^. We must therefore require the scale separation condition

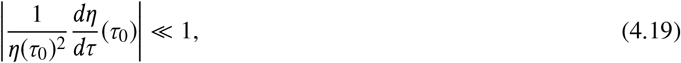

in order for the linear perturbation analysis to make sense. We will check this condition explicitly for each of the perturbation modes turned up by the analysis that follows.

Substituting in Equations (2.2) and (2.3), we find to linear order

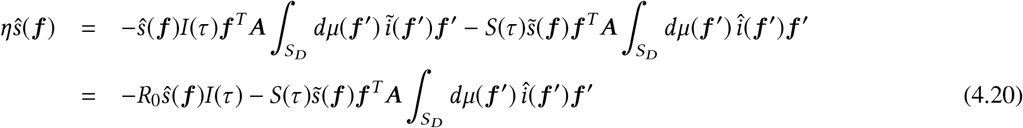

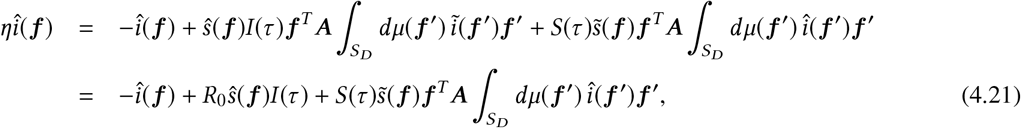

where we have used Equations (3.7) and (3.11) to produce the terms proportional to *R*_0_. Summing these equations, we find again that 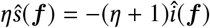.

#### 4.2.1. Modes Decaying at the Removal-Rate

If *η* = 0, we have the trivial solution *ŝ* = *î* = 0. On the other hand, there is a decaying solution *η* = −1, corresponding to *ŝ*(***f***) = 0 and any *î*(***f***) satisfying 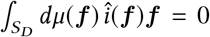. These solutions decay as *e*^−^*^τ^* = e^−^*^νt^*, that is, at the SIR removal rate. Since in this case 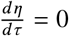, the scale separation assumption of Equation (4.19) holds trivially.

#### 4.2.2. Zero-Moment Modes

Excluding *η* = −1, we again have that

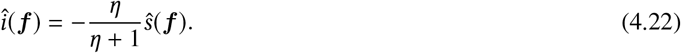

Inserting this relation into Equation (4.20), we can see that if *ŝ*(***f***) satisfies the eigenfunction relation

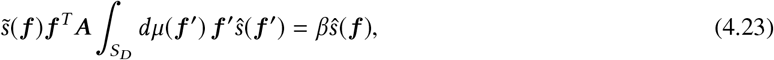

then *η* is a solution of the quadratic equation

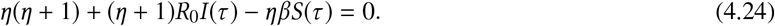

There are two cases to consider. In the first case, the function *ŝ*(***f***) satisfies the zero-moment condition

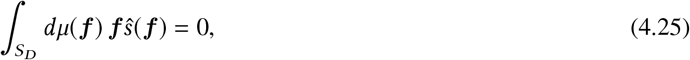

in which case *β* = 0 and by Equation (4.24) we obtain

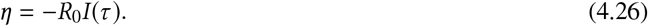

These modes are ostensibly stable. However, at late times, we know from Theorem 3 of Appendix Appendix B that *I*(*τ*) → 0 exponentially. Therefore the ostensible exponential decay time of these modes to the separable solution becomes exponentially large at late times. Unsurprisingly, this occurrence is associated with the late-time failure of the scale separation condition, Equation (4.19):

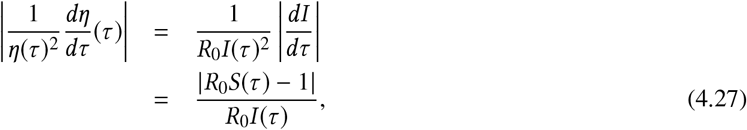

where we have used Equation (3.9) to obtain the last line. We therefore expect scale separation to break down for these modes when

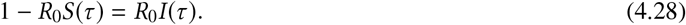

This fact is related to the phenomenon of *freeze-out*, which we discuss in more detail in §4.3.

#### 4.2.3. Finite-Moment Modes

The second case follows the solution procedure for Equation (4.6). From Equation (4.23), the ***f*** -dependence of *ŝ*(***f***) is necessarily of the form

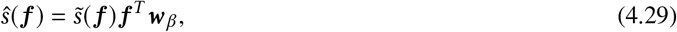

where the *D*-vector ***w****_β_* is independent of ***f***. Inserting this into Equation (4.23), and requiring the result to be true for all ***f***, we find the problem reduced to a *D*-dimensional eigenproblem:

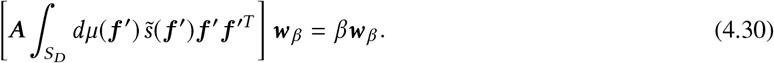

By appealing once again to Theorem 2 in the Appendix, we have that *β* is real and that

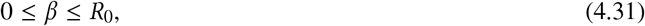

since now Equation (3.11) holds.

One solution to Equation (4.30) is immediate by Equations (3.11) and (2.1): There is an eigenvalue *β* = *R*_0_ corresponding to the right eigenvector ***w****_R_*_o_ = 1 and to the left eigenvector 1^T^ ***A***^−1^. Consequently other ***w****_β_* should be sought in the orthogonal space 1*^T^****A***^−1^ ***w****_β_* = 0. Moreover we learn from this that the upper bound of Inequality (4.31) is sharp, since it holds for at least one eigenvalue. From Equation (4.29), we have that for this eigenmode 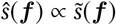.

For each *β*, the solutions to Equation (4.24) are

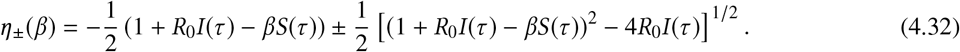

Clearly stability 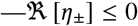—requires that for the largest *β* we have

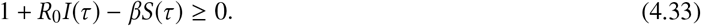

Since we know that at least one eigenvalue achieves the upper bound in Inequality (4.31), the stability condition becomes

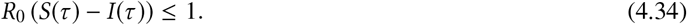

When this condition is achieved, we may expect all deviations from an initial-condition-dependent target separable solution to decrease exponentially at rates *η*_±_(*β*) given by Equation (4.32). We note, however, that as *I*(*τ*) → 0, the *η*_+_ eigenvalues approach zero from below, since

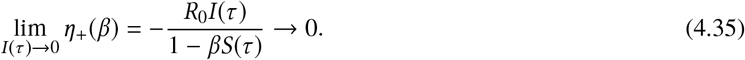

Such modes will not decay to zero rapidly late in the evolution of the epidemic but rather will “freeze out,” as discussed in the next subsection.

### 4.3. Relative Stability of Infection-Free and Separable Solutions

By Part (ii) of Theorem 2, we may rewrite the condition for stability of the infection-free solutions, Equation (4.15), as

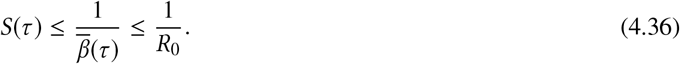

The condition for stability of a separable solution, Equation (4.34), is

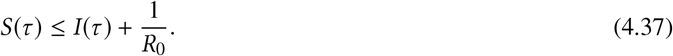

From Equations (4.36–4.37) we see that if a solution initially satisfies neither of the two stability conditions, then the solution will satisfy Equation (4.37) before it satisfies Equation (4.36). It follows that during the initial part of the solution we will have

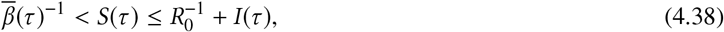

corresponding to a situation in which attraction toward a separable solution begins before attraction toward the infection-free solution sets in. We may therefore infer that a *freeze-out* phenomenon must occur occur, because the eigenvalues of the infection-free solution behave as 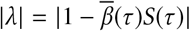, so that their decay rates continue to increase as *S*(*τ*) declines irrespective of *I*(*τ*), whereas the zero-moment and finite-moments modes of the separable solutions have eigenvalues that tend to zero as *I*(*τ*) → 0 according to Equations (4.26) and (4.35). These modes will therefore not efficiently decline toward their target solution at large *τ*, and will be overtaken by the infection-free solution modes.

This phenomenon is evidently related to the fact that the infection-free solutions are globally stable, whereas the separable solutions are not. A related phenomenon is the failure of the scale separation condition for the validity of the perturbation analysis of the separable solutions. Scale separation fails for the moment-free solutions when Equation (4.28) first holds. But if the solution has had time to approach a separable solution, so that 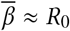, then the left-hand side of Equation (4.28) is approximately the threshold eigenvalue for herd immunity, Equation (4.12), whereas the right-hand side is the eigenvalue of the moment-free modes of the separable solution. Thus loss of scale separation occurs coincidentally with the overtaking of separable mode eigenvalues by infection-free mode eigenvalues, that is, at freeze-out. In fact we may use Equation (4.28) as a rough-and-ready measure of when we may expect freeze-out to occur, since as *I*(*τ*) declines past this point, the eigenvalues X of the infection-free solutions become more negative than a moiety of the eigenvalues of the separable solutions.

We demonstrate the occurrence of the freeze-out phenomenon numerically in what follows.

## 5. Numerical Study

We performed numerical experiments with the BMS system using a *D* = 3 model. We used a triangular discretization on *S*_3_ that is illustrated in Figure 5.1. In each case, the number of subdivisions along each axis is 16, so that there are 256 triangles. After discretization the BMS equations (2.2-2.3) take on the character of a 256-dimensional system of ODEs. Each triangle is ascribed its centroid value of ***f*** as its coordinate for the purpose of computing the source term—the right-hand side of the BMS equations. The integration code is written in Python. We perform the ODE integration using the *scipy.integrate.solve_ivp* method from the *SciPy* package [14]. With the above choice of discretization, all integrations take at most a few seconds running on a laptop computer. Our ternary plot visualizations are made by using *Ternary* [15], a third-party module for the *Matplotlib* [16] Python plotting library.

**Figure 5.1:**
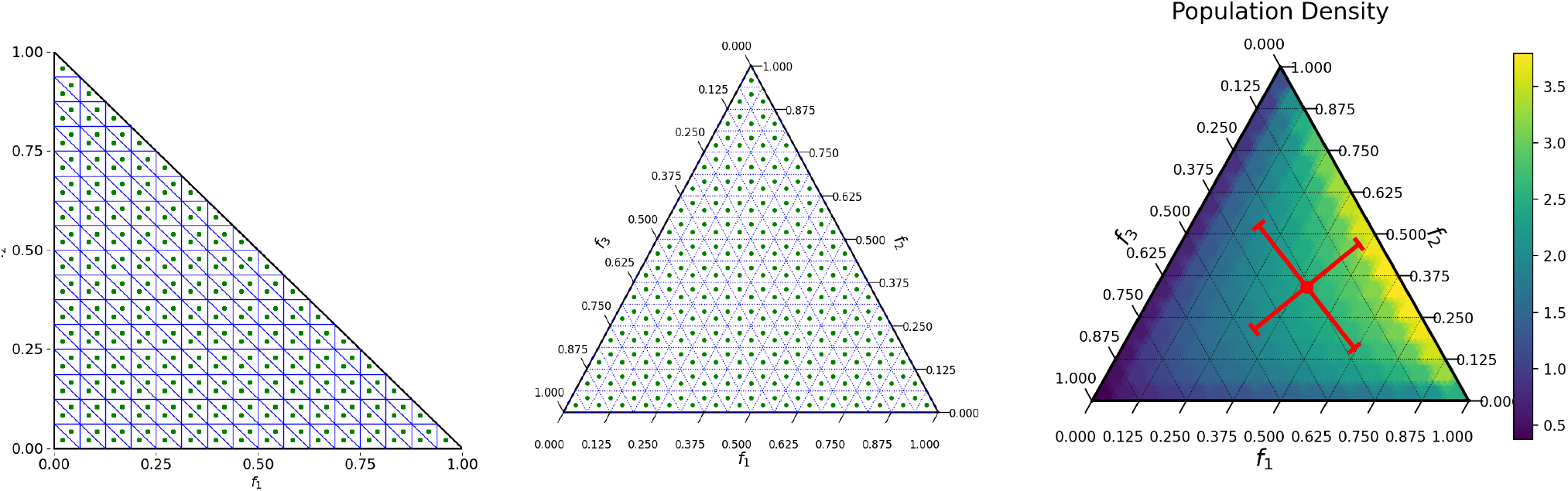
Left panel: Triangularizing discretization used for integration of BMS equations. The figure shows the right triangles in the *f*_1_ − *f*_2_ plane (thin lines) and their centroids (green dots). Middle panel: Ternary plot displaying discretization on the simplex face. The lower-left to upper-right lines are contours of constant *f*_1_, with values shown on the bottom axis. Similarly, *f*_2_ values are displayed on the right axis with left-right contours of constant *f*_2_, and *f*_3_ values are displayed on the left axis with upper-left to lower-right contours of constant *f*_3_. Right panel: Population density on the 3-simplex (Equation 5.1) used for the simulations. The red dot is the distribution centroid. The cross arms represent ±1 standard deviations along the principal covariance directions.

We assume throughout a normalized population density *ρ*(***f***) on the simplex of the Dirichlet type, specifically

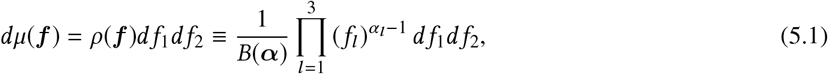

where 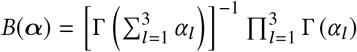 is a normalizing constant. We fix ***α*** = [1.4,1.2,0.9]. The resulting density is shown in the right panel of Figure 5.1. The population-weighted centroid ***f****_pop_*, shown as a red dot in the figure, while the cross shows the principal directions and the root-eigenvalues of the distribution covariance – that is, the cross arms represent ±1 standard deviations along the principal covariance directions. The centroid has the value ***f****_pop_* = [0.398,0.341, 0.260]^t^. Clearly, this density is somewhat biased toward high values of *f*_1_, moderate values of *f*_2_, and low values of *f*_3_. Below we will specify infectivity parameters *a_l_*; such that *l* = 1 corresponds to a low-risk behavior, *l* = 2 corresponds to moderate-low risk, and *l* = 3 corresponds to high-risk superspreader behavior. With these choices, the population is more heavily weighted to lower-risk behaviors 1 and 2 but is by no means free of high-risk behavior 3.

We restore dimensionality to time, choosing a recovery rate *ν* = 0.1 days^−1^, corresponding to an infective period of 10 days.

### 5.1. Epidemic Propagation in Behavior Space

To illustrate the dynamics of epidemic propagation across behavior space—that is, across *S*_3_—we consider an epidemic with parameters *a*_1_ = 25/6, *a*_2_ = 25/3, *a*_3_ = 25, corresponding to a separable solution with *R*_0_ = 2.5 and a centroid ***f****_s_* = *R*_0_***A***^−1^1 = [0.6,0.3, 0.1]^t^. We may interpret these parameters using Equation (2.4), assuming ***f****_s_* = ***f****_i_* = ***f****_pop_*, the population centroid on *S*_3_. The result is that *R_E_* = *R_E_*_, 1_ + *R_E_*_, 2_ + *R_E_*_, 3_ = 0.661 + 0.971 + 1.69 = 3.32, corresponding to a high-risk average 3-behavior, a medium-low-risk average 2-behavior, and a very low-risk average 1-behavior.

We start with a nearly infection-free population, namely, *s*(0, ***f***) = 1, *i*(0, ***f***) = 0 except at location ***f****_i, init_* = [0.95,0.04,0.01] (i.e., near the *f*_1_ = 1 vertex of *S*_3_), where we add to *i*(0, ·) (and subtract from *s*(0, ·)) an amount S*i* corresponding to 10^−4^ of the population, that is, *δi* = 10^−4^/[*ρ*(***f****_i, init_*) × *T*], where *T* is the area of each discretization right triangle in the *f*_1_ – *f*_2_ plane (left panel of Figure 5.1). With this initial choice, the centroids of the initial susceptible and infected distributions are

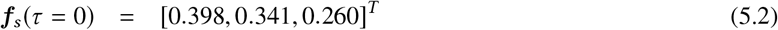

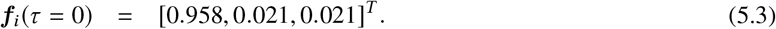

By Equation (2.4) we have *R_E_*(*τ* = 0) = *R_E_*_, 1_ + *R_E_*_, 2_ + *R_E_*_, 3_ = 1.591 + 0.05928 + 0.1355 = 1.79. We can see that the population bias and initial condition have changed the infectivity order, with more initial infectivity ascribed to behavior 1 than to behavior 3.

The left panel of Figure 5.2 displays the progress of the epidemic integrated over *S*_3_. The curves show the profiles of susceptible, infected, and removed populations as a function of time. For comparison we also show the progress of a classic SIR model, started with the same initial infected fraction (10^−4^), with the same v, and with the same *R*_0_ parameter as that corresponding to the separable solution of the BMS model. This figure shows that the BMS model progresses more rapidly than the SIR model. This may seem surprising since the initial *R_Eff_* of the BMS model is 1.79, which is less than the *R*_0_ = 2.5 value used for the SIR model. However, we will see shortly that *R_Eff_* evolves strongly over time and that there is no discrepancy here. The right panel of Figure 5.2 shows the same integrated BMS model, together with vertical dotted lines at the times corresponding to the snapshots of infected fraction distribution on *S_D_* shown in Figure 5.3.

**Figure 5.2:**
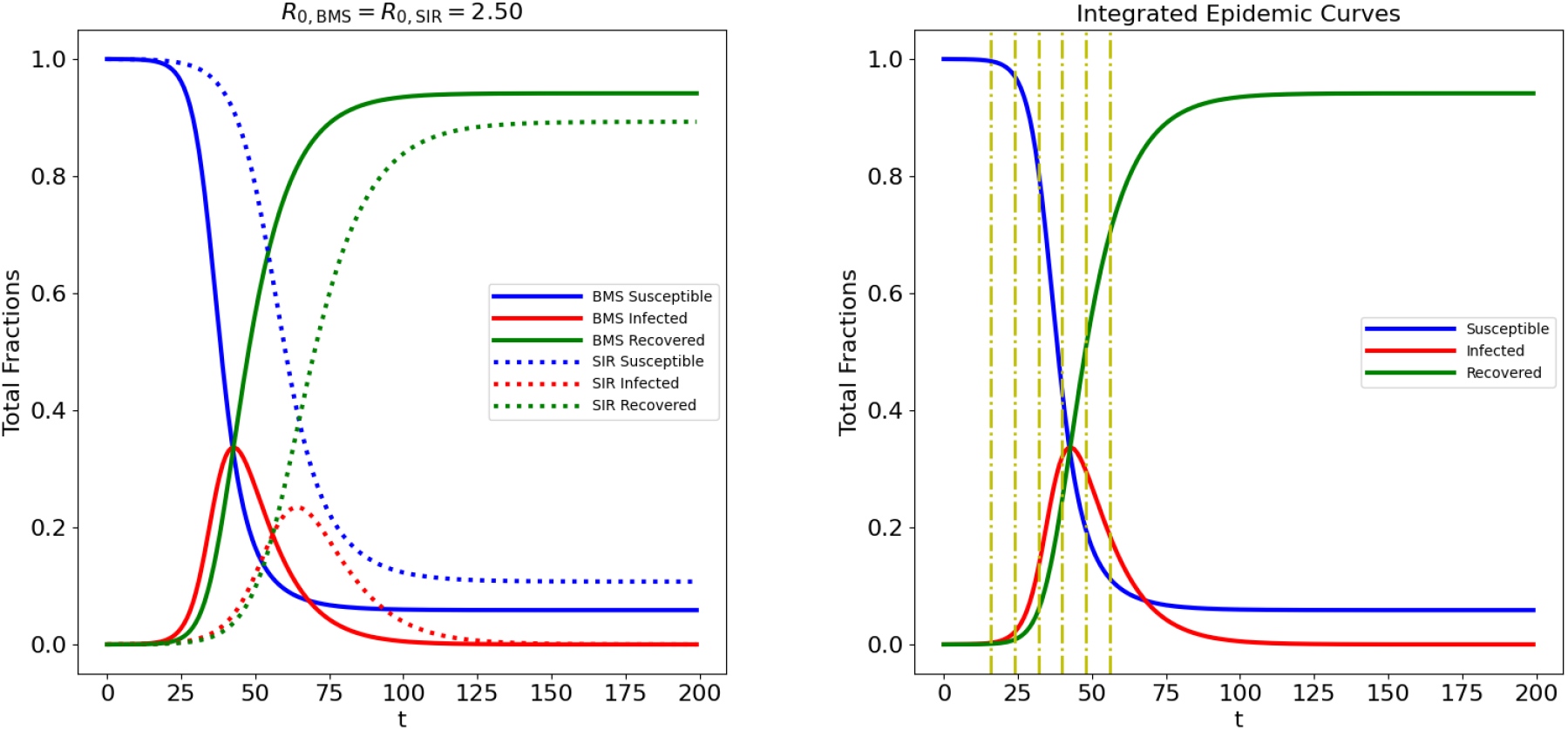
Integrated epidemic progress. Left panel: The solid lines show the susceptible, infected, and removed fractions for the integrated BMS equations initialized as described in §5.1. The dotted lines show the classic SIR model integrated from the same initial state, with the same parameters *R*_0_ and *ν* as the separable-solution parameters of the BMS model. The comparison shows that the BMS model progresses more rapidly than the corresponding SIR model. Right panel: The same BMS model, showing the times at which the snapshots in Figure 5.3 were produced..

**Figure 5.3:**
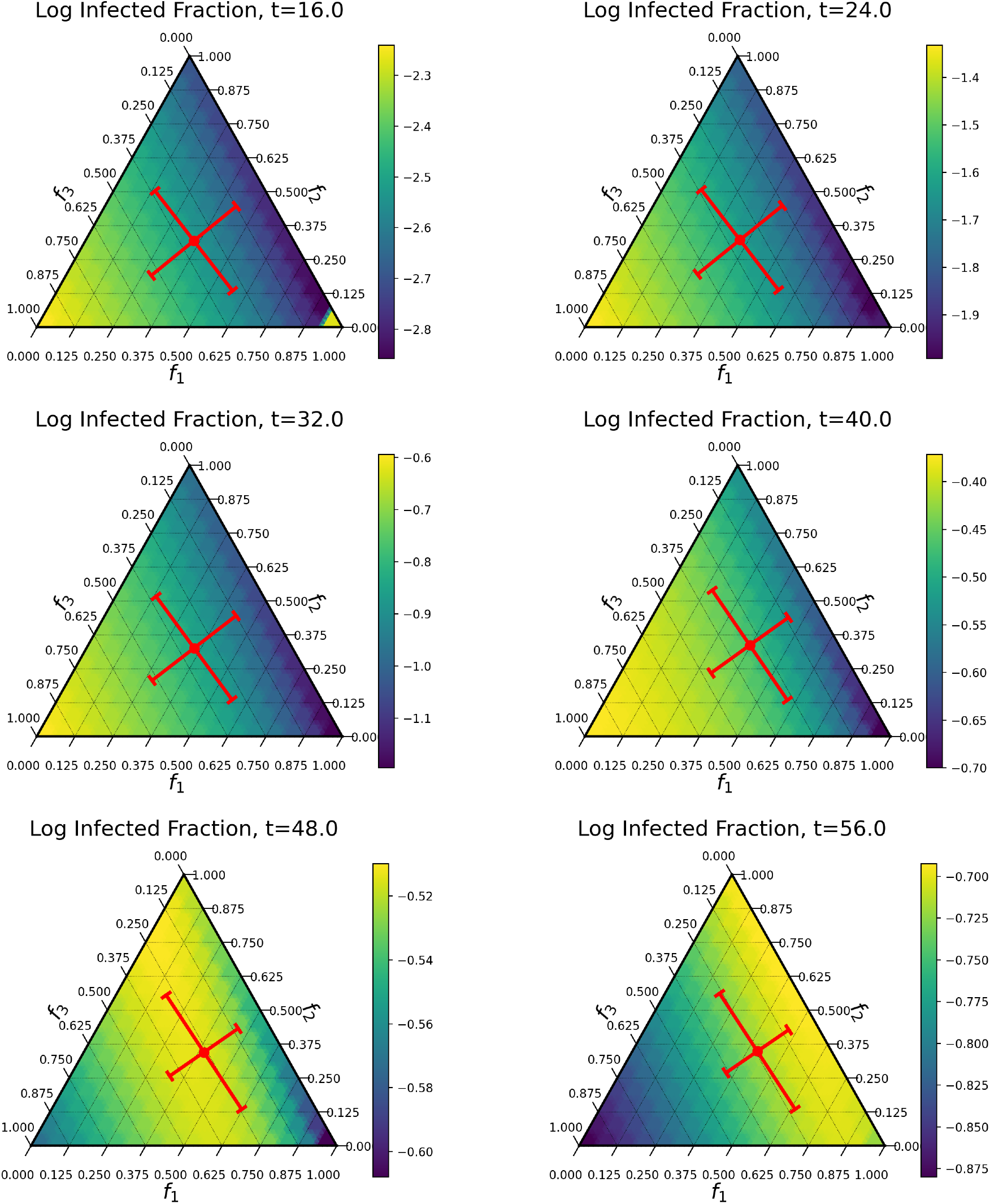
Progress of the epidemic in behavior space, at the times shown in the right panel of Figure 5.2. The panels show infected fraction as a colormap. They also show the centroid and ±1 standard deviation along principal covariance directions of the infectious distribution *dμ*(***f***)*ĩ*(*τ*, ***f***) as a red dot and cross.

The snapshots in Figure 5.3 show infected fraction as a colormap. They also show the centroid and ±1 standard deviation along principal covariance directions of the infectious distribution *dμ*(***f***)*ĩ*(r, ***f***) as a red dot and cross. The first of these snapshots shows the initial infection site at the lower-right corner “reaching over” to infect the high-risk behavior individuals in the lower-left corner. This situation happens rapidly despite the small amount of communication between the two corners in the source term of the BMS equations. Thereafter, we see a wave of infection proceeding from high-risk to low-risk portions of *S*_3_. Animated visualisations of the progress of all BMS variables across behavior space are available in the supplementary materials to this article, as well as at https://mcs.anl.gov/~carlo/Articles/Multispread/.

Figure 5.4 shows the final state of the susceptible population in behavior space. The epidemic has had the greatest effect on the lower-left corner of the space, where the highest-risk behavior is most prevalent. This corner has few remaining susceptibles, compared with the upper-right edge of the diagram, where less risky behavior is prevalent.

**Figure 5.4:**
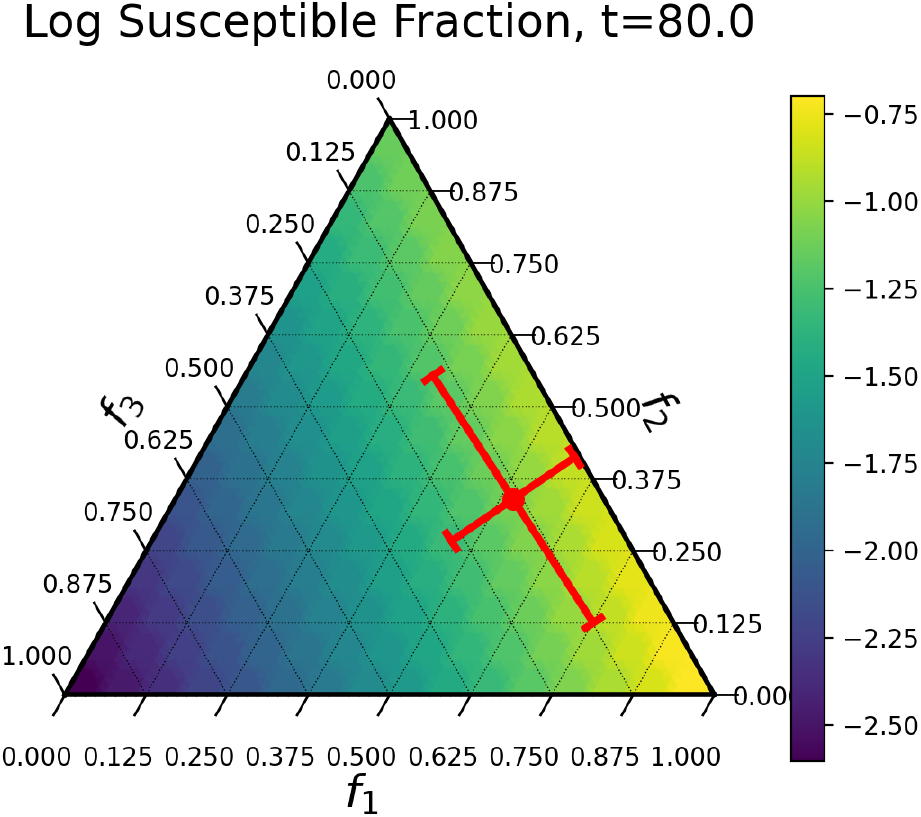
Final state of the susceptible population. The colormap shows the susceptible fraction, while the centroid and ± 1 standard deviation along principal covariance directions of the susceptible distribution d 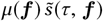 are shown as a red dot and cross. Note that the lower-left corner—the high-risk portion of behavior space—is essentially emptied of susceptibles, whereas susceptibles remain in the lower-risk portion of the space.

### 5.2. Effective Reproduction Numbers and Approach to Separable Solution

Figure 5.5 shows some further properties of the evolution of the BMS equations. The left panel of the figure shows the run of *R_E_*, computed as per Equation (2.4), together with the runs of the three summands of *R_E_*. We see here the resolution of question raised earlier: Why does the BMS solution rise so much more rapidly than the SIR solution, despite starting off with a lower effective reproduction number? As is apparent from the curves, the value of *R_E_* climbs rapidly in the initial phase of the epidemic, driven largely by the contribution to *R_E_* from the high-risk behavior contribution (the red dot-dash line). Thereafter the value of *R_E_* settles down after the epidemic empties the high-risk behavior population of susceptibles. The predicted freeze-out time from Equation (4.28) is shown as a vertical dot-dashed line.

**Figure 5.5:**
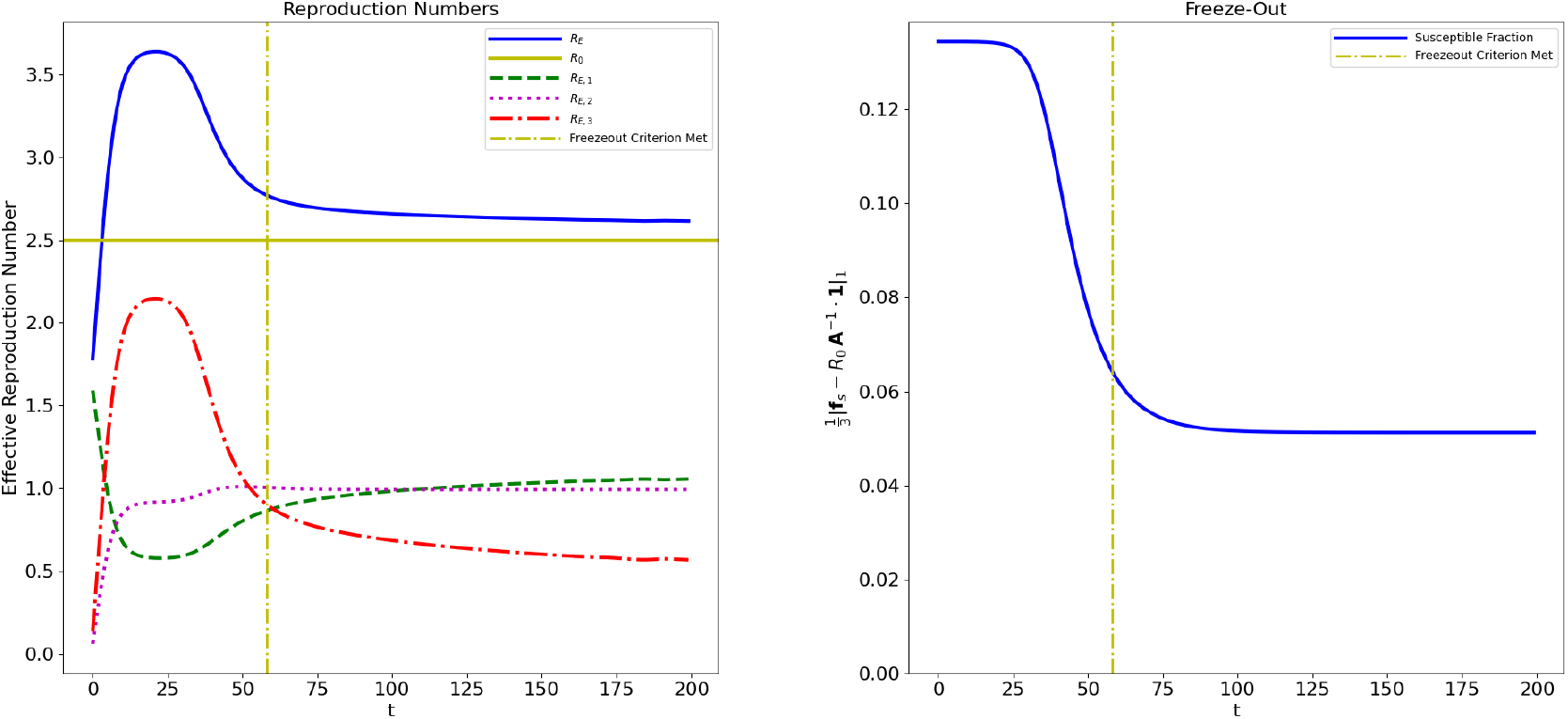
Left panel: *R_E_* and its components. The vertical dot-dashed line shows the time at which the freeze-out criterion (Equation 4.28) is satisfied. The effective reproduction number *R_E_* initially climbs to a high value, driven mostly by the high-risk behavior 3. Eventually *R_E_* settles down, as the epidemic empties the high-risk-behavior region of *S*_3_ of susceptibles. Right panel: Approach to separable solution, measured by the deviation of the mean vector ***f****_s_* from the predicted value for a separable solution (Equation 3.11). The vertical dot-dashed line again shows the predicted freeze-out time, which agrees with the flattening out of the curve.

The right panel of Figure 5.5 illustrates the freeze-out phenomenon. Here we see the average *L*_1_ residual of the mean vector ***f****_s_* from the value predicted for the separable solution by Equation (3.11). We can see that the residual drops rapidly until the predicted freeze-out time, after which it settles toward a steady value.

### 5.3. Other Values of R_0_

Figure 5.6 displays the integrated epidemic progress, effective reproduction numbers, and approach to separable solution for different *R*_0_ parameters—*R*_0_ = 1.0 in the left column and *R*_0_ = 1.5 in the right column. The *a_l_* parameters are simply scaled from their versions for *R*_0_ = 2.5, according to Equation (3.12), preserving the risk ordering of the three behaviors.

**Figure 5.6:**
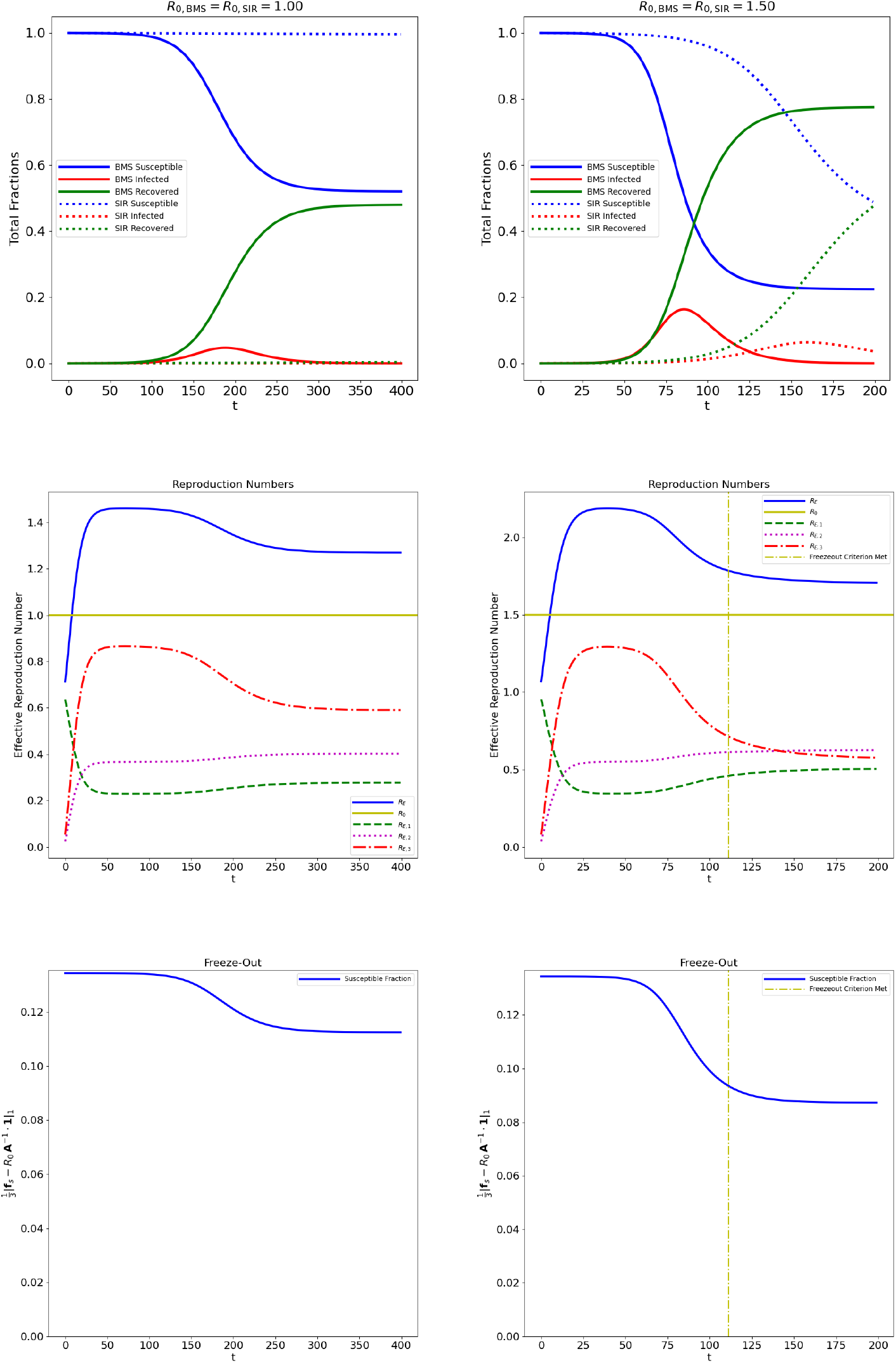
Behavior of BMS model for other values of *R*_0_. Left column: *R*_0_ = 1.0. Right column: *R*_0_ = 1.5. The top row displays epidemic progress integrated over *S*_3_. The middle column shows effective reproduction numbers. The bottom column shows approach to separable solution. The yellow dot-dashed line showing the advent of freeze-out is not displayed for *R*_0_ = 1.0 since the freeze-out condition of Equation (4.28) is never attained in this case.

In the *R*_0_ = 1 case, we observe that there is an epidemic notwithstanding the fact that the corresponding separable case—SIR with *R*_0_ = 1-is dormant, as shown in the top figure. In fact, the system state never progresses far enough toward its separable solution to see an appreciable decline in effective reproduction number from the initial value, as is apparent in the middle panel. The lower panel again shows anemic progress toward the separable solution. In fact, in this case the freeze-out condition of Equation (4.28) is never attained.

The *R*_0_ = 1.5 case resembles the *R*_0_ = 2.5 case, although less progress occurs toward the separable solution.

### 5.4. An Actual Separable Solution

Given the importance of separable solutions to the theory presented in this work, an interesting verification is to obtain an actual separable solution numerically. We may obtain such a solution by the following simple expedient. Run a series of integrations of the BMS equations. At each stage, initialize the normalized shape functions 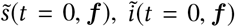 to the final corresponding shape functions from the previous iteration, and reset the integrated variables *S*(*t* = 0) = 1 – 10^−4^, *I*(*t* = 0) = 10^−4^. This procedure allows the convergence to a separable solution to restart and proceed further before freeze-out sets in again. The results of four iterations of this process using the same parameters and initial condition as in §5.1 are displayed in Figure 5.7. The left panel shows that the integrated quantities *S*(*t*), *I*(*t*), R(t) are now indistinguishable from the corresponding quantities in the corresponding SIR model, as expected. The middle panel displays the effective reproduction numbers, which are nearly constant and which add up to *R*_0_ = 2.5, as expected. The right panel displays the final log normalized population density of the susceptibles, 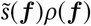, where *ρ*(***f***) is the population density with respect to the measure *df*_1_*df*_2_ defined in §2.1. This distribution shows a pronounced tilt toward the low-risk-behavior sector that accords well with intuition about the progress of an epidemic.

**Figure 5.7:**
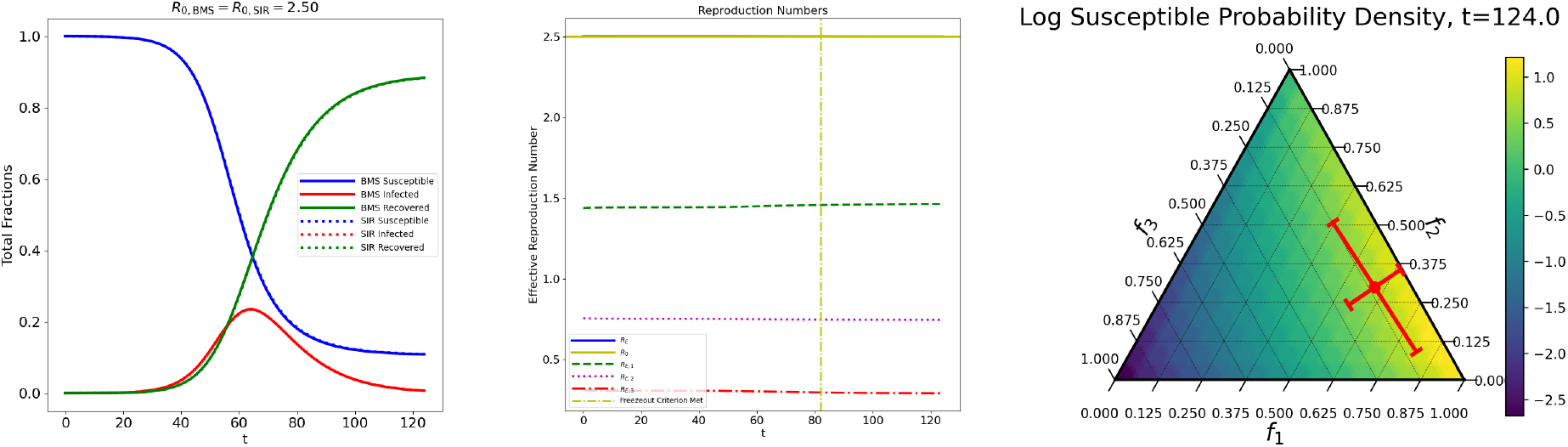
Separable solution, obtained by iterating 4 times the process of initializing each integration using the final normalized shape functions 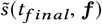, *ĩ*(*t_final_*, ***f***) from the previous iteration, and resetting the integrated parameters *S*(*t* = 0) = 1 – 10^−4^, *I*(*t* = 0) = 10^−4^. The left panel shows the integrated variables, which are now indistinguishable from their SIR counterparts. The middle panel shows effective reproduction numbers, which are essentially constant and which sum to R_0_ = 2.5. The right panel shows the final population distribution density of susceptibles in behavior space, which shows a pronounced tilt in favor of the low-risk-behavior sector.

## 6. Discussion

We have presented a new dynamical system, the behavioral multispread model, motivated by the desirability of extending compartmental models to behavior space so as to create a context where the behavioral usage of the term “superspreader” has a technical-mathematical meaning, analogous to that corresponding to the individual-physiology and event usages of the term. In this work, we have largely focused on the mathematical properties of the BMS system, with particular regard to solution stability and long-term behavior, as well as comparisons with the ancestor of BMS, the SIR system. We demonstrated that the BMS model is, unsurprisingly, capable of exhibiting richer dynamical behavior than is SIR, with dynamically evolving effective reproduction numbers and a more complex description of herd immunity. It is particularly interesting to observe the evolution of *R_E_*, since this represents transitions in the susceptible and infected distributions in behavior space. As these distributions change, the relative importance of the infectivities corresponding to different behaviors also change.

We now discuss the status of BMS in the field of mathematical epidemic models and sketch some possible directions for future research.

An unexplored mathematical topic in connection with the BMS model is to answer the questions “What happens when *D* gets very large, or even infinite?” Does there exist some useful limit, and if so, what are its properties?

At this stage, BMS is useful principally as an illustrative model of epidemic propagation in behavior space, rather than as a detailed model capable of comparison with epidemic data, let alone detailed prediction. In order to make BMS into an epidemiological model capable of realistic description and interpretation, some obvious research steps need to take place. In the first instance, it may be that *D* = 3 is too simplistic a representation of behavior space. In fact there is likely an infinitude of risk behaviors, but it is not implausible that they could be grouped into a few coarse categories. After all, a behavior such as bar-hopping may well have a parameter *a_l_* that is close to that of another behavior such as theater-going, so that the two activities look indistinguishable from the model’s point of view, and the only difference between our hypothetical populations of bar-hoppers and theater-goers is that their respective distributions in behavior space differ, because they spend different amounts of their time at these (and other) activities.

Once a reasonable set of risk-activity categories is determined, a second problem is determining the population density *ρ*(***f***) over the corresponding behavior space. This is evidently a sociological challenge, possibly to be addressed using data from surveys, censuses, and the like.

A technical issue that can arise is one of visualization. For this work, we chose *D* = 3 because visualization of 3-component compositions on the 3-simplex is elegantly accomplished via ternary diagrams. Higher-dimensional visualizations would be more challenging and might be better accomplished by other types of diagrams.

Another issue is discretization. The 3-simplex *S*_3_ lends itself to a straighforward triangular decomposition, which can be understood and coded based on simple Euclidean geometry. Higher-dimensional “triangularizations” require input from more advanced computational geometry. A separate but related issue is stability of numerical integration. As carried out in this work, the integro-differential equations of the BMS model were summarily discretized, thus trading the MBS system in for a (large) system of ODEs. Fortunately for us, this worked flawlessly, and in the cases we examined we ran into no troublesome issues such as frequently attend naive discretizations. However, in this work we can offer no guarantees that this should be so irrespective of discretization resolution or system dimensionality. A study of the numerical properties of BMS would thus be of some value.

We point out that despite the aforementioned difficulties in using BMS as a realistic, interpretable model, there is no difficulty in its use as an empirical fitting model, and potentially some advantage to using it in this way. Given the rich dynamical behavior of BMS compared, say, with SIR, the ease with which the system can be numerically discretized, and the efficiency and speed of integration, there could be value in fitting parameters such as *α_l_*;, *a_l_*; to the progress of a real epidemic, assuming sufficient data is available, to investigate whether, for example, the BMS characteristic of a rapidly increasing and then settling-down effective reproduction number can provide a better description of certain types of epidemics.

## Data Availability

Animations of model solutions are available in supplementary materials, as well as at the URL below.

https://www.mcs.anl.gov/~carlo/Articles/Multispread/

## Acknowledgments

This material is based upon work supported by the U.S. Department of Energy, Office of Science, under contract number DE-AC02-06CH11357.

## Appendix A. Eigenvalues of Stability Matrix

We now give proof of the assertions made in §4 concerning the eigenvalues of the *D*-dimensional stability matrix 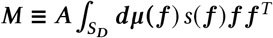. We first establish a necessary lemma.

### Lemma 1

*Fix* ***h*** ∊ *S_D_ and* ***w*** ∊ *ℝ^D^. Let* 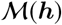 *be the space of probability measures dη*(***f***) *on S_D_ satisfying* 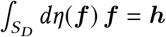*. Define the objective function O*: 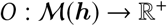 *by*

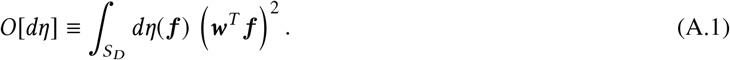

*Then the solution of the maximization problem*

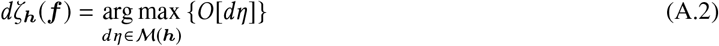

*is the (****w****-independent) additive superposition of Dirac-δ measures supported at the simplex vertices given by*

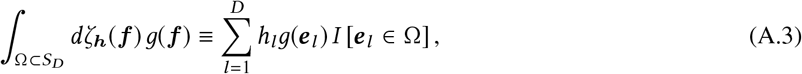

*where the e_l_ are unit vectors in the l-direction, in other words*, [*e_l_*]*_m_* = *δ_lm_, and I* [] *is the indicator function*.

It is straightforward to establish that *dζ_h_*(***f***) is a probability measure that satisfies the mean constraint, since *dζ_h_*(***f***) is manifestly non-negative definite, 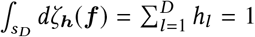 since ***h*** ∊ *S_D_*, and 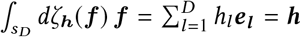. For the measure *dζ_h_*(***f***) the objective of Equation (A.1) becomes 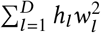. We will establish the lemma by proving that any finite perturbation of *dζ_h_*(***f***) that preserves the mean constraint leads to a diminution of the objective function from this value.

We define the perturbed measure *dη*(***f***) = *dζ_h_*(***f***) + *dγ*(***f***), where the measure *dγ*(***f***) may be an additive mixture of a measure that is absolutely continuous with respect to the reference measure *dμ*(***f***) (i.e., admitting a Radon-Nikodym density function) and some singular measures such as Dirac-*δ* components, and including Dirac-*δ* components at the simplex vertices that adjust the corresponding components of *dζ****_h_f***). In other words,

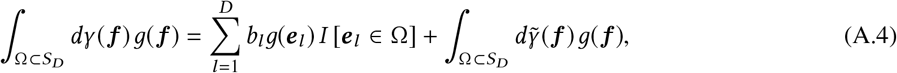

where the non-negative-definite measure 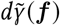 has support excluding the simplex vertices and may itself be an additive superposition of absolutely continuous and singular measures.

Imposing the mean constraint, we have

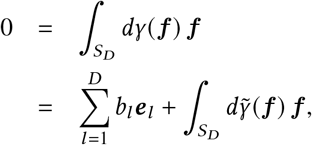

whence

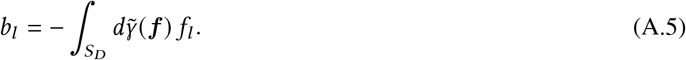

The perturbation to the objective function of Equation (A.2) is therefore

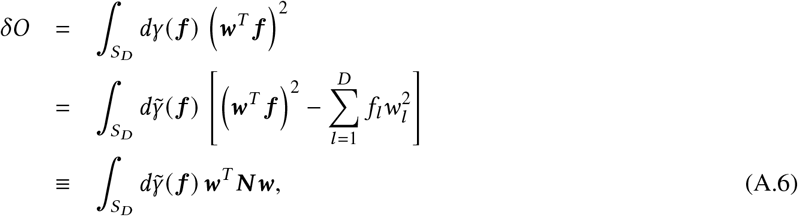

where we have defined the symmetric matrix ***N*** with components [***N***]*_lm_* = *f_l_ f_m_* – *f_l_δ_lm_*. Since the measure 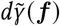 is non-negative definite, we may establish the lemma by proving that the eigenvalues of ***N*** are all negative or zero.

So consider the eigenvector equation ***Nr*** = *α****r***, which is to say

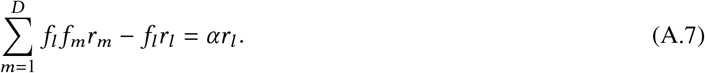

Since ***f*** ∊ *S_D_*, it follows that ***r*** = **1** is an eigenvector with eigenvalue *α* = 0. One also can easily show that ***f*** is not an eigenvector of ***N***. We may therefore choose the normalization ***f****^T^****r*** = 1 for ***r***. Equation (A.7) then becomes

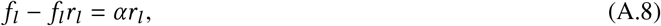

from which we obtain

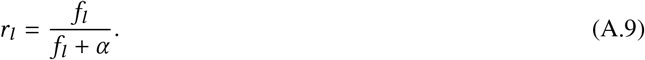

Imposing the normalization condition ***f****^T^* **r** = 1, we obtain the eigenvalue equation

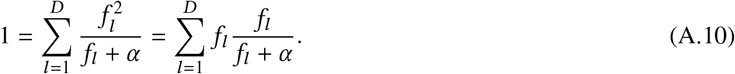

If *α* > 0, however, we have that *f_l_*/[*f_l_* + *α*] < 1 for all *l*, so

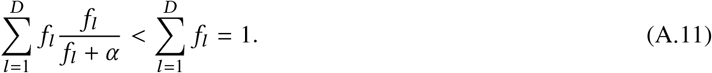

This would contradict Equation (A.10). It follows that the eigenvalues of ***N*** satisfy *α* ≤ 0. Therefore *δO* ≤ 0 in Equation (A.6), and the lemma is proven. □

A corollary of the lemma that we do not require but that is nonetheless interesting is that the covariance matrix 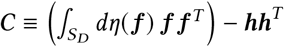 has variances ***w****^T^C****w*** that are maximal when *dη*(***f***) = *dζ****_h_***(***f***).

We may now use the lemma to demonstrate the required statement about the eigenvalues of the stability matrix of the BMS equations.

### Theorem 2

*Suppose that* ***A*** *is a positive-definite diagonal D-dimensional matrix Diag*(*a*_1_,… *a_D_*), *and that* 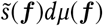 *is a probability measure on S_D_ with first moment* 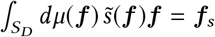*. Then:*

i. *The eigenvalues β of the stability matrix* 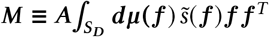 *are real and satisfy* 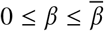*, where*

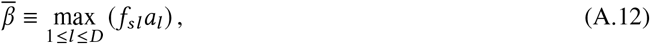

*and where f_sl_ is the l-th component of* ***f****_s_*.
ii. 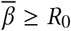.
iii. *If A*, 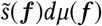*, and R*_0_ *are related as in Equation (3.11), so that f_sl_* = *R*_0_/*a_l_*, *then* 0 ≤ *β* < *R*_0_.

To prove reality and non-negativity, we observe that if ***Mx*** = *β****x***, then

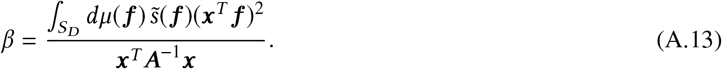

This expression is real and non-negative, because of the positive-definiteness of ***A***.

Now set ***y*** = ***A*** ^1/2^**x** and 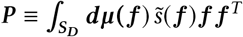. Then we have *β* = *g*(***y***), where

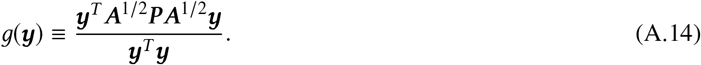

By the Courant-Fisher minimax theorem [17, p.441] the function *g*(***y***) is maximal when ***y*** is the eigenvector of ***A***^1/2^***PA***^1/2^ corresponding to the largest eigenvalue. But ***A***^1/2^***PA***^1/2^ = ***A***^−1/2^***MA***^1/2^, so that ***M*** and ***A***^1/2^***PA***^1/2^ are related by a similarity transformation. ***M*** and ***A***^1/2^***PA***^1/2^ therefore have the same eigenvalues, and their respective eigenvectors x and y are related by ***A***^−1/2^***x*** = ***y***.

We therefore have that if *β_max_* is the maximum eigenvalue of ***M***, then it is also the maximum eigenvalue of ***A***^1/2^***PA***^1/2^, so that

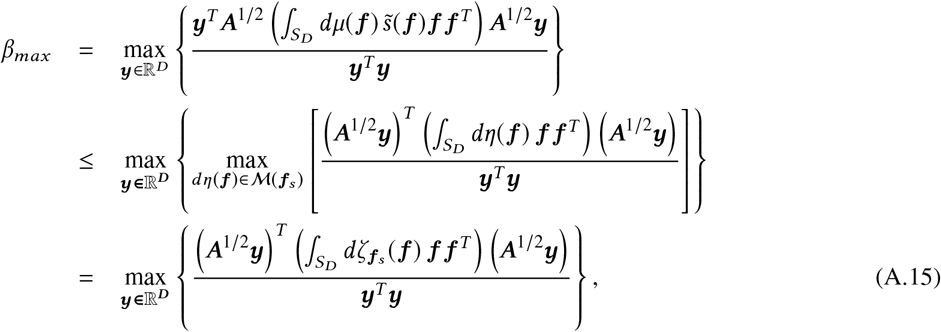

where we have used Lemma 1 to obtain the last line and *dζ****_f_****_s_*(***f***) is the singular measure in Equation (A.3). We have

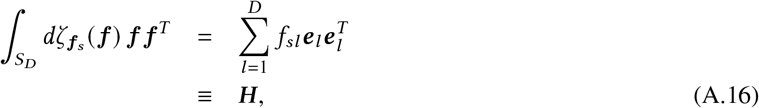

where ***H*** = Diag(*f_s_*_1_,…, *f_sD_*). Combining Equations (A.15) and (A.16), we obtain

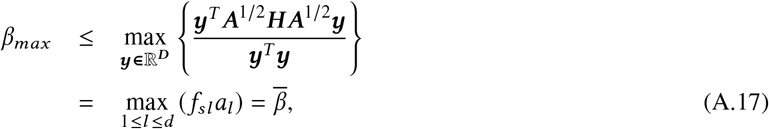

since ***A*** and ***H*** are both diagonal matrices. This completes the proof of (i).

To prove (ii), we note that 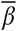 is minimized when all terms *f_sl_a_l_* are equal to each other. Indeed, suppose that *f_sm_a_m_* is larger than the remaining *f_sl_a_l_*, *l* ≠ *m*. Then we may decrease 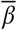 by decreasing *f_sm_* while increasing other components *f_sl_* to maintain the simplex condition **1***^T^* ***f****_s_ =* 1. On the other hand, if all the *f_sl_a_l_* are equal, then any perturbation of ***f****_s_* preserving the simplex constraint will necessarily increase at least one of the *f_sl_a_l_*, resulting in an increase in 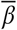.

Hence the minimum of 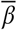 occurs when *f_sl_a_l_* = *c*, so that *f_sl_ = c*/*a*_l_. By the simplex constraint, 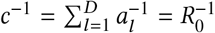. It follows that 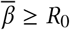.

To prove (iii), observe that when Equation (3.11) holds, we have *f_sl_ = R_0_/a_l_*, and

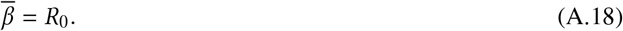

## Appendix B. Global Stability of Infection-Free Solutions

We now prove a result on the global stability of the infection-free solutions of the BMS equations.

### Theorem 3

*Any solution s* (*τ*, ***f***), *i* (*τ*, ***f***) *of the BMS equations with a total susceptible fraction* 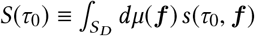 *satisfying the herd immunity condition at time τ*_0_,

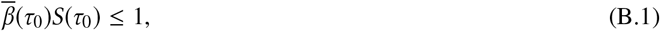

*where* 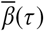 *is given by Equation (4.14), necessarily converges to an infection-free solution as τ* → *∞*.

The evolution of the total infected fraction *I*(*τ*) is given by Equation (2.6),

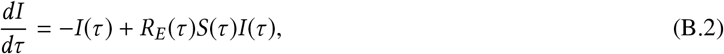

with *R_E_*(*τ*) given by

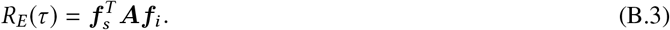

The distribution moments ***f****_s_*, ***f****_i_* are given in Equations (2.7-2.8).

Assume that *S*(*τ*_0_) satisfies the condition of Equation (B.1). The quantity 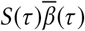 may be expanded by combining Equations (4.14) and (2.7):

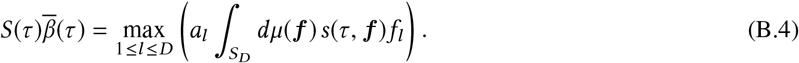

Differentiating each term in the argument of the max function on the right-hand side of this equation with respect to *τ*, we may observe that

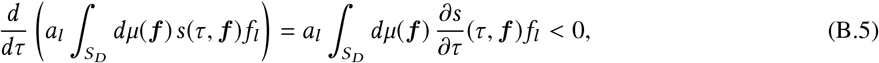

since by Equation (2.2) the integrand term 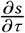 is strictly negative for non-zero *s*(*τ*, ***f***) and *i*(*τ*, ***f***). It follows that all the terms in the argument of the max function in the right-hand side of Equation (B.4) decline with time, and that after a finite time Δ*τ* there will exist a finite positive constant c such that

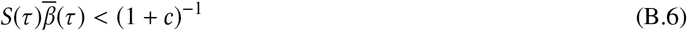

for *τ* > *τ*_0_ Δ*τ*.

Combining this with Equations (B.6) and (B.2) we find that for *τ* > *τ*_0_ + Δ*τ*,

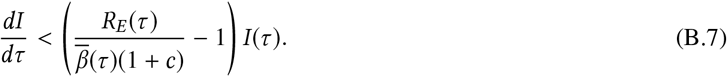

Substituting for *R_E_*(*τ*) from Equation (B.3) and for 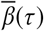 from Equation (4.11) this becomes

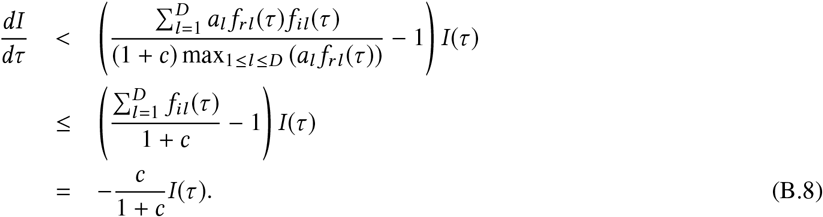

Integrating this inequality with respect to *τ*, we obtain

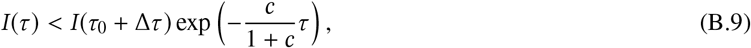

which vanishes in the limit *τ* → 0. Hence in the asymptotic limit such solutions are always infection-free. □

## Government License

The submitted manuscript has been created by UChicago Argonne, LLC, Operator of Argonne National Laboratory (“Argonne”). Argonne, a U.S. Department of Energy Office of Science laboratory, is operated under Contract No. DE-AC02-06CH11357. The U.S. Government retains for itself, and others acting on its behalf, a paid-up nonexclusive, irrevocable worldwide license in said article to reproduce, prepare derivative works, distribute copies to the public, and perform publicly and display publicly, by or on behalf of the Government. The Department of Energy will provide public access to these results of federally sponsored research in accordance with the DOE Public Access Plan. http://energy.gov/downloads/doe-public-access-plan.

